# The relevance of rich club regions for functional outcome post-stroke is enhanced in women

**DOI:** 10.1101/2022.06.28.22277020

**Authors:** Anna K. Bonkhoff, Markus D. Schirmer, Martin Bretzner, Sungmin Hong, Robert W. Regenhardt, Kathleen L. Donahue, Marco J. Nardin, Adrian V. Dalca, Anne-Katrin Giese, Mark R. Etherton, Brandon L. Hancock, Steven J. T. Mocking, Elissa C. McIntosh, John Attia, John W. Cole, Amanda Donatti, Christoph J. Griessenauer, Laura Heitsch, Lukas Holmegaard, Katarina Jood, Jordi Jimenez-Conde, Steven J. Kittner, Robin Lemmens, Christopher R. Levi, Caitrin W. McDonough, James F. Meschia, Chia-Ling Phuah, Stefan Ropele, Jonathan Rosand, Jaume Roquer, Tatjana Rundek, Ralph L. Sacco, Reinhold Schmidt, Pankaj Sharma, Agnieszka Slowik, Alessandro Sousa, Tara M. Stanne, Daniel Strbian, Turgut Tatlisumak, Vincent Thijs, Achala Vagal, Johan Wasselius, Daniel Woo, Ramin Zand, Patrick F. McArdle, Bradford B. Worrall, Christina Jern, Arne G. Lindgren, Jane Maguire, Ona Wu, Natalia S. Rost, the MRI-GENIE and GISCOME Investigators and the International Stroke Genetics Consortium

**Affiliations:** J. Philip Kistler Stroke Research Center, Massachusetts General Hospital, Harvard Medical School, Boston; Univ. Lille, Inserm, CHU Lille, U1171 – LilNCog (JPARC) – Lille Neurosciences & Cognition, F-59000, Lille, France; Computer Science and Artificial Intelligence Lab, Massachusetts Institute of Technology, Boston, USA; Athinoula A. Martinos Center for Biomedical Imaging, Department of Radiology, Massachusetts General Hospital, Charlestown, MA, USA; Department of Neurology, University Medical Center Hamburgl_JEppendorf, Hamburg, Germany; Johns Hopkins School of Medicine, Department of Psychiatry, Baltimore, MD; Hunter Medical Research Institute, Newcastle, New South Wales, Australia; School of Medicine and Public Health, University of Newcastle, NSW, Australia; Department of Neurology, University of Maryland School of Medicine and Veterans Affairs Maryland Health Care System, Baltimore, MD, USA; School of Medical Sciences, University of Campinas (UNICAMP) and the Brazilian Institute of Neuroscience and Neurotechnology (BRAINN), Campinas, SP, Brazil; Department of Neurosurgery, Geisinger, Danville, PA, USA; Research Institute of Neurointervention, Paracelsus Medical University, Salzburg, Austria; Department of Emergency Medicine, Washington University School of Medicine, St Louis, MO, USA; Department of Neurology, Washington University School of Medicine & Barnes-Jewish Hospital, St Louis, MO, USA; Department of Clinical Neuroscience, Institute of Neuroscience and Physiology, Sahlgrenska Academy, University of Gothenburg, Sweden; Department of Neurology, Sahlgrenska University Hospital, Gothenburg, Sweden; Department of Neurology, Neurovascular Research Group (NEUVAS), IMIM-Hospital del Mar (Institut Hospital del Mar d’Investigacions Mèdiques), Department of Medicine and Life Sciences (MELIS)- Universitat Pompeu Fabra, Barcelona, Spain; KU Leuven - University of Leuven, Department of Neurosciences, Experimental Neurology and Leuven Research Institute for Neuroscience and Disease (LIND), Leuven, Belgium; VIB, Vesalius Research Center, Laboratory of Neurobiology, University Hospitals Leuven, Department of Neurology, Leuven, Belgium; School of Medicine and Public Health, University of Newcastle, Newcastle, New South Wales, Australia; Department of Neurology, John Hunter Hospital, Newcastle, NSW, Australia; Department of Pharmacotherapy and Translational Research and Center for Pharmacogenomics, University of Florida, Gainesville, FL, USA; Department of Neurology, Mayo Clinic, Jacksonville, FL, USA; Department of Neurology, Clinical Division of Neurogeriatrics, Medical University Graz, Graz, Austria; Henry and Allison McCance Center for Brain Health, Massachusetts General Hospital, Boston, MA, USA; Department of Neurology and Evelyn F. McKnight Brain Institute, Miller School of Medicine, University of Miami, Miami, FL, USA; Institute of Cardiovascular Research, Royal Holloway University of London (ICR2UL), Egham, UK & St Peter’s and Ashford Hospitals, UK; Department of Neurology, Jagiellonian University Medical College, Krakow, Poland; Department of Laboratory Medicine, Institute of Biomedicine, the Sahlgrenska Academy, University of Gothenburg, Gothenburg, Sweden; Department of Neurology, Helsinki University Hospital and University of Helsinki, Helsinki, Finland; Stroke Division, Florey Institute of Neuroscience and Mental Health, Heidelberg, Australia and Department of Neurology, Austin Health, Heidelberg, Australia; Department of Radiology, University of Cincinnati College of Medicine, Cincinnati, OH, USA; Department of Clinical Sciences Lund, Radiology, Lund University; Department of Radiology, Neuroradiology, Skåne University Hospital, Lund, Sweden; Department of Neurology and Rehabilitation Medicine, University of Cincinnati College of Medicine, Cincinnati, OH, USA; Department of Neurology, Pennsylvania State University, Hershey, PA, USA; Division of Endocrinology, Diabetes and Nutrition, Department of Medicine, University of Maryland School of Medicine, Baltimore, MD, USA; Departments of Neurology and Public Health Sciences, University of Virginia, Charlottesville, VA, USA; Department of Clinical Genetics and Genomics, Sahlgrenska University Hospital, Gothenburg, Sweden; Department of Neurology, Skåne University Hospital, Lund, Sweden; Department of Clinical Sciences Lund, Neurology, Lund University, Lund, Sweden; University of Technology Sydney, Sydney, Australia

**Keywords:** Bayesian hierarchical modeling, lesion-symptom mapping, rich club, functional outcome, sex differences

## Abstract

This study aimed to investigate the influence of stroke lesions in pre-defined highly interconnected (rich club) brain regions on functional outcome post-stroke, determine their spatial specificity and explore the effects of biological sex on their relevance.

We analyzed MRI data recorded at index stroke and ∼3-months modified Rankin Scale (mRS) data from patients with acute ischemic stroke (AIS) enrolled in the multisite MRI-GENIE study. Structural stroke lesions were spatially normalized and parcellated into 108 atlas-defined bilateral (sub)cortical brain regions. Unfavorable outcome (mRS>2) was modeled in a Bayesian logistic regression framework that relied on both lesion location, as well as the covariates: age, sex, total DWI lesion volume and comorbidities. Effects of individual brain regions were captured as two compound effects for (i) six bilateral rich club and (ii) all further non-rich club regions. Via model comparisons, we first tested whether the rich club region model was superior to a baseline model considering clinical covariates and lesion volume only. In spatial specificity analyses, we randomized the split into “rich club” and “non-rich club” regions and compared the effect of the actual rich club regions to the distribution of effects from 1,000 combinations of six random regions. In sex-specific analyses, we introduced an additional hierarchical level in our model structure to compare male and female-specific rich club region effects.

A total of 822 patients (age: 64.7 (standard deviation: 15.0), 39% women, 27.7% with mRS>2) were analyzed. The rich club model substantially outperformed the baseline model (weights of model comparison: rich club model: 0.96; baseline: 0.04). Rich club regions had substantial relevance in explaining unfavorable functional outcome (mean of posterior distribution: 0.08, area under the curve: 0.8). In particular, the rich club-combination had a higher relevance than 98.4% of random constellations (15/1,000 random constellations with higher mean posterior values). Among the these 15 random constellations with higher means, the most frequently selected regions were the inferior temporal gyrus (posterior division, 8/15), the putamen (8/15), the cingulate gyrus (7/15) and the superior parietal lobule (6/15). Rich club regions were substantially more important in explaining long-term outcome in women than in men (mean of the difference distribution:-0.107, 90%-HDPI:-0.193 to -0.0124).

Lesions in rich club regions were associated with increased odds of unfavorable outcome. These effects were spatially specific, i.e., the majority of random combinations of six regions had comparably smaller effects on long-term outcome. Effects were substantially more pronounced in women.

## Introduction

Stroke is the most burdensome neurological disorder in the US, surpassing both Alzheimer disease and migraine with respect to absolute Disease-Adjusted Life Years (DALYs).^1^ Enhancing our understanding of underlying factors of severe disease is a stepping stone in designing tailored acute and rehabilitative stroke therapies and improving stroke outcome in the longer term.^2,3^

One particularly promising avenue, when aiming to understand the neurobiological effects of ischemic stroke lesions, is to build upon our current understanding of physiological brain organization – with its network structure as a core element.^4,5^ Specific to this network conceptualization is the assumption of a hub structure, i.e., highly interconnected brain regions constituting the so-called rich club.^6^ These rich club brain regions are assumed to form the backbone for functional integration of diverse brain networks and, hence, large-scale, inter-regional communication.^7^ In their seminal study involving healthy participants, van den Heuvel and Sporns identified six bilateral regions central to this rich club: Superior parietal and frontal lobules, precuneus, thalamus, putamen and hippocampus.^6^ Subsequent studies suggest joint genetic underpinnings,^8^ higher metabolic needs^9^ and critical implications in cognition of these rich club regions.^10^ What is more, neuropsychiatric diseases, such as Alzheimer’s dementia,^11^ schizophrenia^12^ and epilepsy,^13^ show a tendency to affect these rich club regions primarily. This later observation constitutes the “nodal stress hypothesis”.^11,14^

Recent work in the stroke field has also adopted the notion of stroke as a network disease^15^ and started to integrate connectome-derived information in stroke outcome models. In particular, these approaches successfully established links between stroke lesions in highly central brain regions and functional outcomes,^16,17^ cognitive functions,^18,19^ aphasia^20^ and motor recovery post-stroke.^21^ However, it is important to appreciate that these studies primarily tested whether brain regions central to the network structure, i.e., *a priori* defined brain regions, had the capacity to explain outcome. In most cases, these studies did not assess any spatial specificity aspects, that is whether their *a priori* chosen constellations of brain regions were indeed more informative than random constellations of brain regions.

The current study aimed to complement previous approaches by scrutinizing the spatial specificity of classically assumed rich club regions in their relevance for functional stroke outcome in a broad, unselected multi-center stroke sample. To this end, we designed a probabilistic lesion-symptom mapping framework and employed permutation analyses to probe the rich club constellation against 1,000 random constellations of brain regions. Leveraging our Bayesian models’ flexibility, we also examined whether there were any sex-related differences in the relevance of rich club regions. We hypothesized that rich club regions would have a disproportionately important role in explaining stroke outcome and recovery. In view of our previous findings of enhanced effects of left-hemispherical posterior circulation lesions in women,^22,23^ with many of these posterior regions being part of the rich club, we furthermore hypothesized that we would find more augmented rich club effects in women compared to men.

## Methods

### Ischemic stroke patient cohort

In this complete case study, we included all MRI–Genetics Interface Exploration (MRI-GENIE) patients with AIS,^24^ that had readily available, high-quality DWI-derived lesion segmentations,^25^ clinical information on sociodemographic/clinical characteristics (age, sex, comorbidities), and follow-up modified Rankin Scale (mRS) data (c.f., **supplementary materials** for a sample size calculation). We employed the same inclusion and exclusion criteria as in our previous work.^23^ It is important to note that the research questions and analytical approaches of our previous and this current work differ substantially as outlined below. The results presented here are hence novel. All subjects gave written informed consent in accordance with the Declaration of Helsinki. The study protocol was approved by Massachusetts General Hospital’s Institutional Review Board (Protocol #: 2001P001186 and 2003P000836) and Review Boards of individual sites.

### Sociodemographic, clinical and neuroimaging data

We considered age, sex, available cardiovascular risk factors (hypertension (HTN), coronary artery disease (CAD), diabetes mellitus (DM), atrial fibrillation (AF), history of smoking and prior stroke), and the mRS-derived functional outcome (7-point score: 0=no detectable symptoms, 2=slight disbility, 3=moderate disability, 6=death). Data on age, sex and comorbidities was acquired during hospitalization, and functional outcome was recorded between day 60 to day 190 post-stroke. Neuroimaging scans, more specifically diffusion-weighted images (DWI), were collected during the acute hospital stay (in the majority of cases in the first 48h, c.f., **supplementary materials** for a description of imaging parameters). DWI-derived stroke lesion segmentations were generated via a previously validated ensemble of 3-dimensional convolutional neural networks.^25^ We non-linearly normalized DWI images and respective DWI-lesion segmentations to Montreal Neurological Institute (MNI)-space. Results underwent careful quality control by two experienced raters (A.K.B., M.B.) to ensure a high quality of lesion segmentation and spatial normalization. We then calculated the number of stroke lesion-affected voxels per atlas-defined area in 94 cortical and 14 subcortical bilateral brain regions (the unilateral brainstem parcel was excluded, as non-hemisphere-specific region).^26^ These 108 brain regions were divided into “rich-club” and “non-rich-club” regions according to previous work by van den Heuvel and Sporns.^6^ The rich club here consisted of bilateral cortical precuneus, superior frontal and superior parietal cortex parcels, as well as subcortical bilateral hippocampus, putamen and thalamus parcels (**Figure 1**). The definition and structural extent of rich club regions did not differ between men and women.

**Figure 1.**
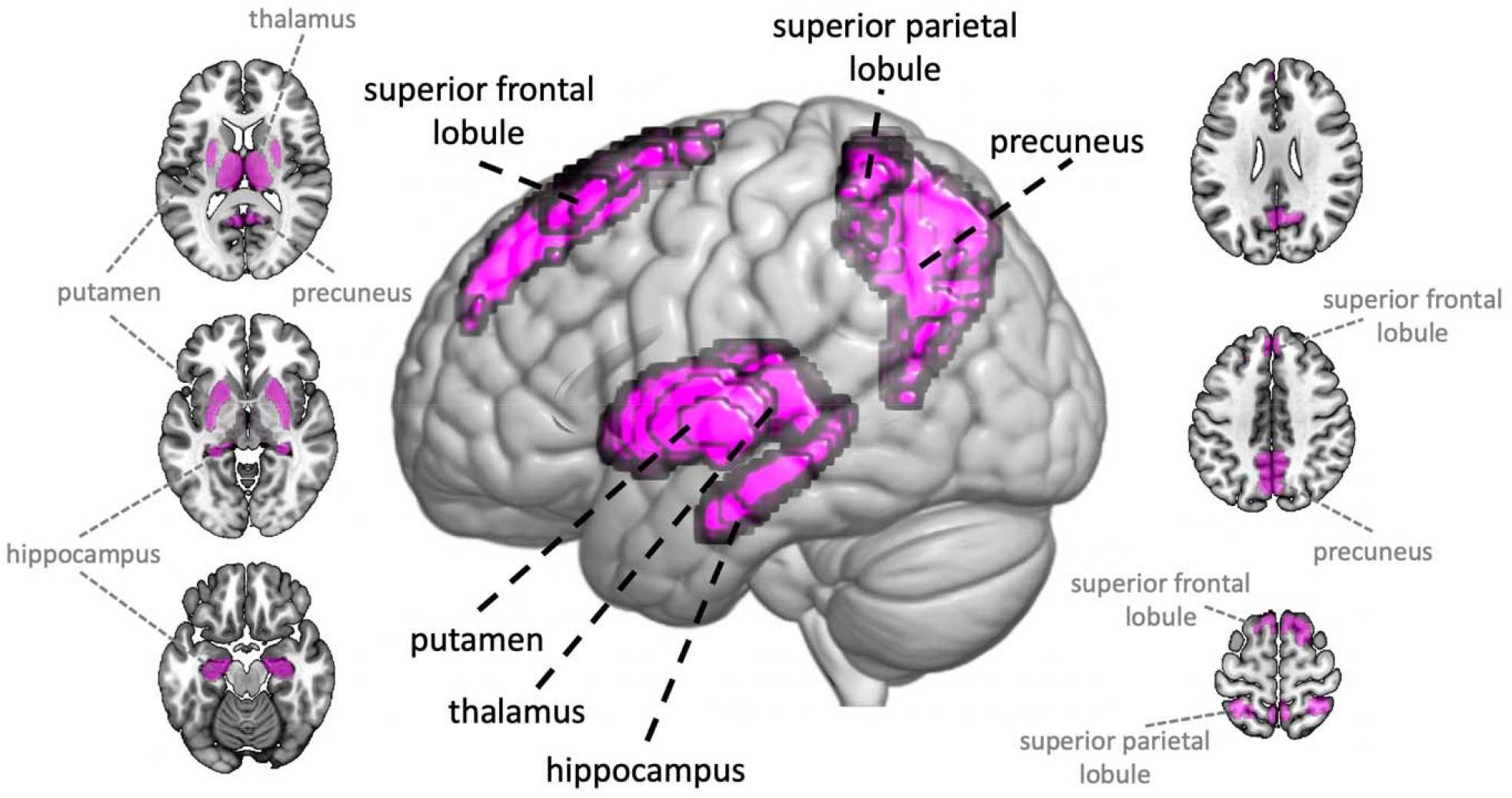
Brain renderings of rich club regions, as defined in work by van den Heuvel and Sporns.^6^. In the present study, we focused on six bilateral brain regions: The superior parietal and frontale lobules, the precuneus, the thalamus, the putamen and the hippocampus.

### Modeling unfavorable functional outcomes

We employed Bayesian logistic regression to model unfavorable outcome (mRS>2).^27^ Brain region-specific lesion effects were captured separately for “rich club” and “non-rich club” brain regions within a hierarchical model structure, i.e., we designed two hyperparameters on the higher level that summarized the effects of the six bilateral “rich club” and 48 bilateral “non-rich club” regions. In addition to the lesion information, we accounted for (mean-centered) age, age^2^, sex, total lesion volume, and the presence of following known cardiovascular risk factors: hypertension, diabetes mellitus type 2, atrial fibrillation, coronary artery disease, prior stroke and smoking. Covariates were chosen line with previous work.^22,23^ Both age and age^2^ were included to correct for linear, as well as non-linear U-shaped age effects (e.g., if the outcome is affected the same way in both younger and older, but not middle-aged patients). As in previous work,^23^ we refrained from including initial stroke severity as a covariate, as it conceivably represents the extent and location of brain injury. The full model specifications are stated in our **supplementary materials**.

Samples were drawn from the Bayesian posterior parameter distributions via the No U-Turn Sampler (NUTS), a type of Monte Carlo Markov Chain algorithm (setting: draws=2500).^28^ The model performance was evaluated as the area under the curve (AUC). While we refrained from interpreting individual region-wise effects in view of the higher dimensional input space, we focused on interpreting collapsed rich club and non-rich club effects.

### Comparison to the baseline model

To ensure that information on lesion location, as captured in our atlas-defined ROIs, substantially augmented outcome prediction performance, we first conducted a Bayesian model comparison with a baseline model. This baseline model considered clinical characteristics and total DWI stroke lesion volume only.

### Permutation analysis

Our main aim was to estimate the overall effect of lesions to rich club regions on unfavorable outcome post-stroke. Accordingly, the model parameter of interest was the hyperprior ***mu_*** β_***rich club***_ summarizing all individual rich club region effects. We evaluated the sampled Bayesian posterior distribution of ***mu_*** β_***rich club***_ several ways. First, we compared the overall rich club region effect to the overall non-rich club region effect by subtracting both of their posterior distributions, similar to our previous work.^22,23,29,30^ We defined substantial differences as 90% highest probability density intervals (HDPIs) of difference distributions not overlapping with zero. Furthermore, we conducted permutation analyses: In these analyses, we randomly selected six bilateral brain regions and combined them as competing “rich club” regions. The non-selected brain regions were subsequently designated “non-rich club” regions. We then ran the same logistic regression model, as described for the main analyses. This step was repeated 1,000 times.

We computed the mean values of all 1,000 sampled posterior distributions for random “rich club” combinations and compared the resulting distribution of mean values to the mean value determined for the original “real” rich club combination. In particular, we determined the number of mean values higher or equal to the real rich club regions’ mean value. We counted how often each region was selected to gain insight into which brain regions contributed to constellations resulting in comparably high or higher mean values than for the “real” rich club constellation.

### Sex-specific rich club effects

Further analyses centered on sex-specific effects of lesions to the rich club. As in previous analyses,^22^ we integrated hyperprior(s) that were capable of capturing overall rich club and non-rich club effects separately for men and women. We then evaluated differences between hierarchically estimated female and male-specific rich club and non-rich club effects via contrasting of the corresponding posterior parameter distributions. Lastly, we tested whether there were any significant sex differences in either the total or parcel-wise lesion volumes and the frequencies with which each parcel was affected (p<0.05, FEW-corrected).

### Data and code availability

The authors agree to make the data available to any researcher for the express purposes of reproducing the here presented results and with the explicit permission for data sharing by individual sites’ institutional review boards. The Harvard-Oxford atlas can be downloaded here: https://fsl.fmrib.ox.ac.uk/fsl/fslwiki/Atlases. Bayesian analyses were implemented in Python 3.7 (predominantly relying on packages: nilearn^31^ and pymc3^32^).

## Results

### Stroke patient characteristics

We included a total of 822 patients with AIS in this study (mean age: 64.7 (15.0), 39.2% women). Favorable 3-months outcomes (mRS<3) were achieved by 72.3% of all patients, the median score on the modified Rankin Scale was 1 (interquartile range (IQR): 0-3). The cohort was furthermore characterized by 64.1% patients with HT, 21.8% with DM, 16.8% with AF, 18.4% with CAD, 9.7% with prior stroke and 55.0% with smoking as cardiovascular risk factors. An exhaustive display of patients’ characteristics, differentiating between men and women, is shown in **Table 1**. The maximum overlay of structural stroke lesions was found to be subcortically, surrounding the lateral ventricles. Lesions were equally distributed between the left and right hemispheres and showed similar spatial distributions for men and women (**Figure 2**).

**Table 1.** Patient characteristics. Mean values and standard deviation, unless otherwise noted. Characteristics of men and women were compared either via two-sample t-tests or two-sided Fisher’s exact tests as appropriate.

|  | **All participants**<br>**(n=822)** | **Male**<br>**participants**<br>**(n=500)** | **Female**<br>**participants**<br>**(n=332)** | **Statistical**<br>**comparison**<br>**of male and**<br>**female**<br>**participants** |
| --- | --- | --- | --- | --- |
| **Age** | 64.7 (15.0) | 63.9 (14.2) | 65.8 (16.2) | p=0.07 |
| **Female Sex** | 39.2% | - | - | - |
| **Acute stroke severity**<br>**(median,**<br>**interquartile range)** | 4 (5) | 4 (5) | 4 (6) | p=0.07 |
| **3-months mRS**<br>**(median,**<br>**interquartile range)** | 1 (0-3) | 1 (1-2) | 2 (0-3) | p<0.001 |
| **Normalized DWI-**<br>**derived stroke lesion**<br>**volume (ml, median,**<br>**interquartile range)** | 3.3 (0-12.8) | 2.9 (0-11.3) | 3.8 (0-17.8) | p=0.28 |
| **Hypertension** | 64.1% | 63.0% | 65.9% | p=0.41 |
| **Diabetes mellitus**<br>**type 2** | 21.8% | 23.0% | 19.9% | p=0.30 |
| **Atrial fibrillation** | 16.8% | 14.6% | 20.2% | p=0.04 |
| **Coronary artery**<br>**disease** | 18.4% | 21.8% | 13.0% | p=0.002 |
| **Smoking** | 55.0% | 61.0% | 45.7% | p<0.001 |
| Prior stroke | 9.7% | 9.4% | 10.2% | p=0.72 |

**Figure 2.**
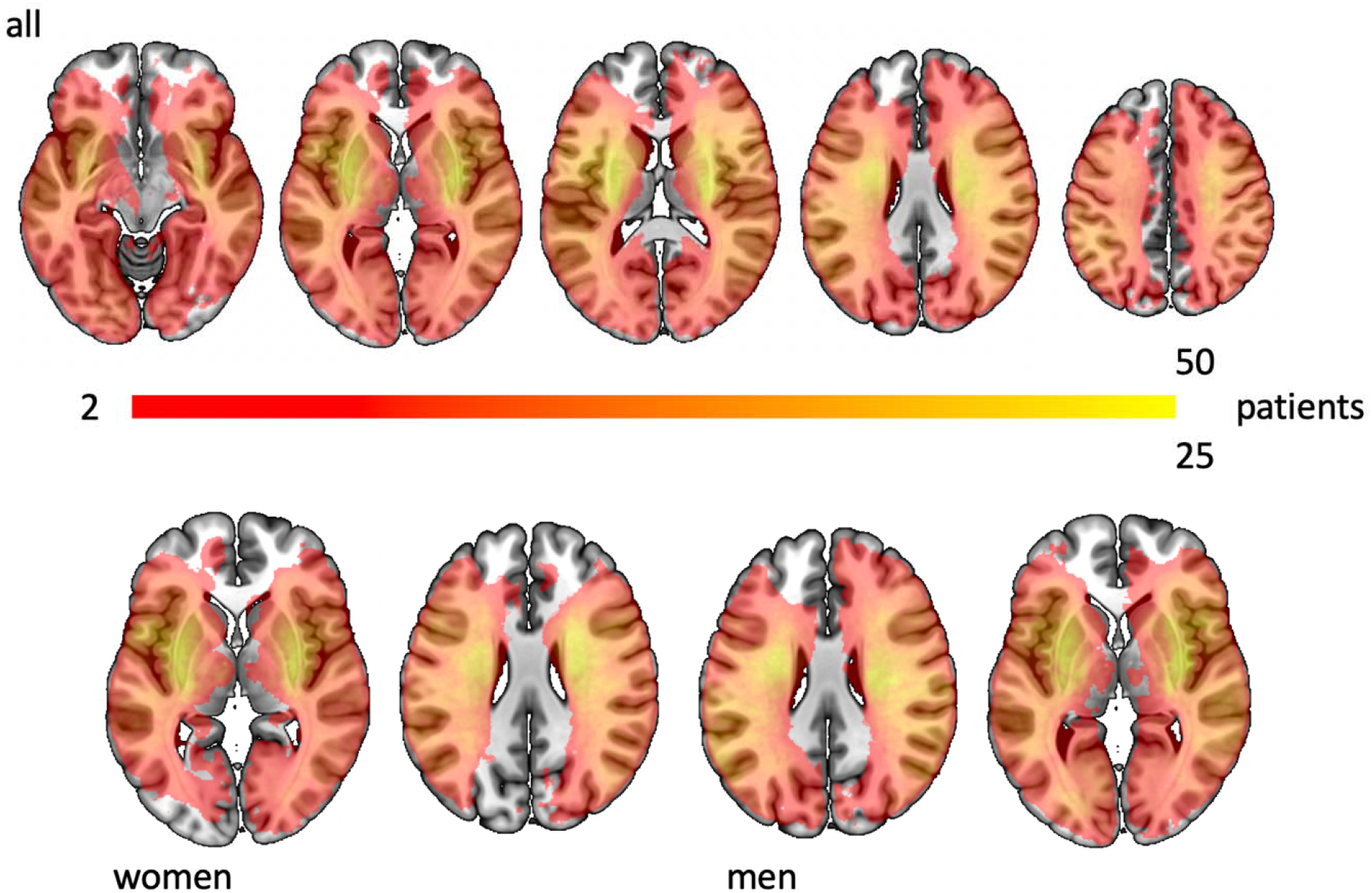
Lesion overlaps of all patients (*upper row*) and specifically for all female and male patients (*bottom row*).

### Prediction of unfavorable functional outcomes

The AUC for modeling unfavorable outcome (mRS>2) by relying on our main rich club model was 0.80. As demonstrated by a leave-one-out cross validation-based model comparison, the rich club model noticeably outperformed the baseline model that considered information on total lesion volume, but not individual lesion location. Consequently, the rich club model was considered superior (model weights assigned during model comparison: rich club model: 0.96; baseline: 0.04, **Supplementary Figure 1**).

The overall rich club region hyperprior effect indicated increased odds of unfavorable outcomes in case of rich club region lesion (posterior mean: 0.08, 90%-HPDI: 0.04 to 0.13, **Figure 3**, *upper left*). In particular, this overall rich club region effect was substantially larger than the respective one for all other non-rich club brain regions combined (difference in posterior mean: -0.08, 90%-HDPI: -0.13 to -0.03). The covariates age, female sex, DM, prior stroke and total DWI lesion volume all independently increased the odds of unfavorable outcomes (age: posterior mean: 0.04, 90%-HPDI: 0.03 to 0.06; female sex: posterior mean: 0.42, 90%-HPDI: 0.15 to 0.74; DM: posterior mean: 0.645, 90%%-HPDI: 0.29 to 0.99; prior stroke: posterior mean: 1.4, 90%-HPDI: 0.94 to 1.8; lesion volume: posterior mean: 0.19, 90%-HPDI: 0.08 to 0.3). Further covariates (HTN, CAD, AF, Smoking) were not associated (**Figure 3**). What is more, our ancillary analyses indicated that the rich club effect, as apparent in the main analyses, could predominantly be traced back to women: The compound rich club effect was substantially more pronounced in women compared to men, as suggested by the difference in Bayesian posterior distributions that did not overlap with zero (difference in posterior distributions: mean: -0.107, 90%-HDPI: -0.193 to -0.0124). At the same time, we did not observe any statistically significant differences in total or parcel-wise lesion volumes or parcel-wise lesion status frequency (all *p*>0.05, FWE-corrected).

**Figure 3.**
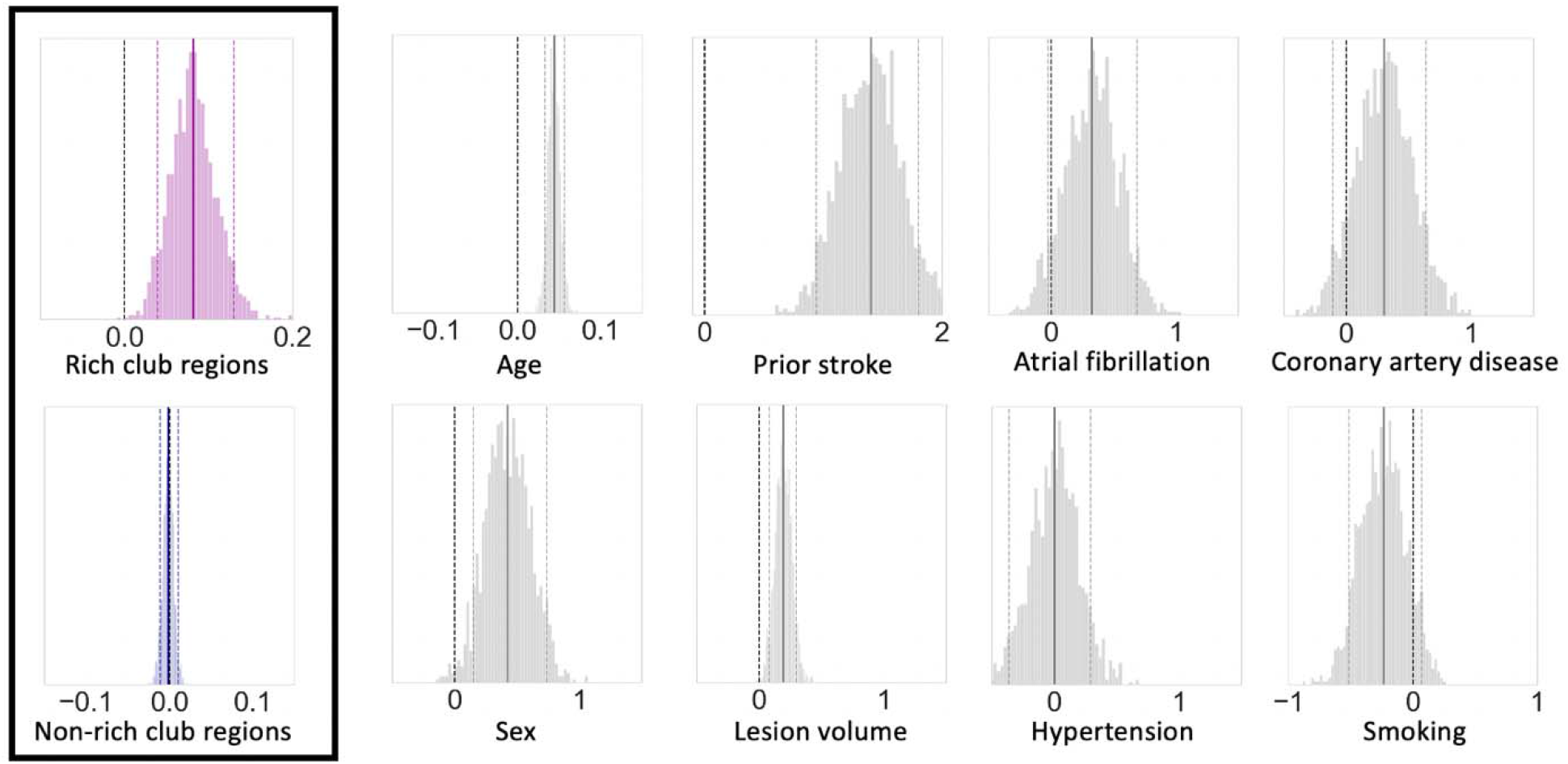
Bayesian posterior distributions of the non-sex-specific rich club region model. We considered effects to be substantially relevant, if the 90% highest probability density intervals, as marked by the dashed lines, did not overlap with zero. Correspondingly, lesions in rich club regions, as well as the covariates age, female sex, DM, prior stroke and total DWI lesion volume were all positively associated with increased odds of unfavorable ∼3-months outcome. Additionally, the rich club region effect emerged as substantially more pronounced than the non-rich club region effect in direct comparisons of posterior distributions.

### Specificity analyses

The overall effect of rich club regions was greater than the effects of 98.4% of the random brain region combinations. In absolute numbers, only 15 out of the 1,000 random constellations exceeded the effect of the original rich club combination. The combination of regions achieving the highest effect comprised the superior parietal lobule, the subcallosal cortex, the occipital fusiform gyrus, the superior temporal gyrus (anterior division), and the inferior temporal gyrus (anterior and posterior division**; Figure 4**). The inferior temporal gyrus (posterior division), the putamen (both in 8 out of the 15 constellations), the cingulate gyrus (7/15) and the superior parietal lobule (6/15) were the most frequently involved parcels in these 15 constellations. Altogether, the majority (13) of these 15 constellations included at least one rich-club region. The remaining two constellations interestingly overlapped in encompassing parcels relating to the cingulate gyrus, the inferior temporal gyrus and the fusiform gyrus, suggesting their potential importance in functional outcome modeling.

**Figure 4.**
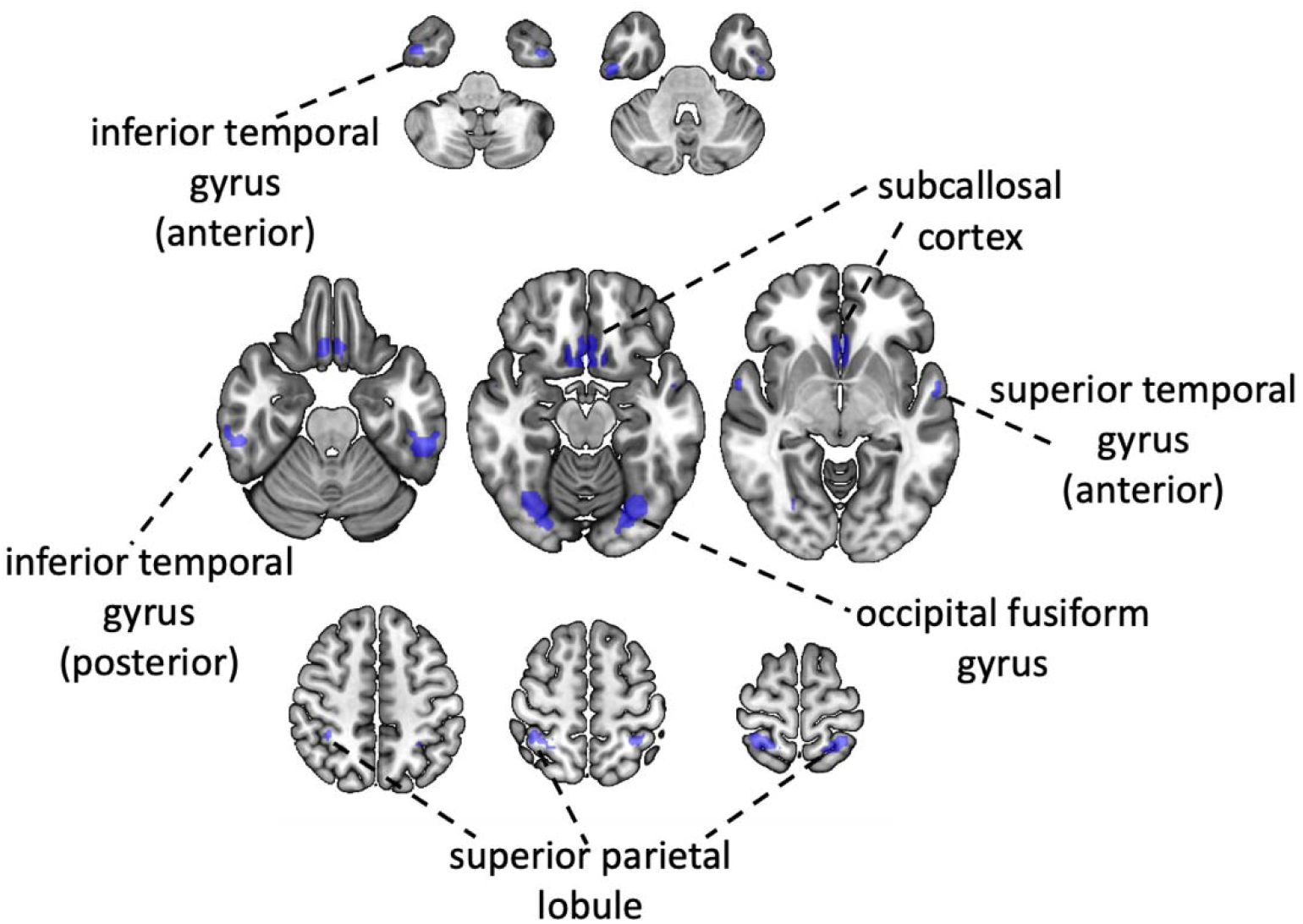
Visualization of bilateral brain regions constituting the combination of random “rich club” regions with the highest overall effect on ∼3-months functional outcomes post-stroke. The superior parietal lobule was the only region being part of both the real rich club constellation, as well as random combination. While there was no contribution from subcortical regions in this best performing random combination, further regions were distributed all across the cortical surface, i.e., featuring frontal, temporal, parietal and occipital regions. Temporal regions were the most frequently represented ones.

## Discussion

Rich club regions represent critical nodes for cerebral information transfer.^6^ Capitalizing on a large, multi-site international sample of patients with AIS, we show that this combination of structurally-defined rich club regions has a prime effect on outcome in the subacute phase post-stroke. Permutation analyses exemplified that the rich club combination exceeded the effects on outcomes of 98.4% of random brain region combinations. In sex-specific analyses, biological female sex emerged as a potential key driver of this rich club effect.

### Rich club-focused lesion studies now and then

The critical prime role of rich club regions as determined here, is well in line with previous studies: These studies carved out hub region involvement in a manifold of neuropsychiatric diseases,^14^ such as Alzheimer’s dementia,^11^ schizophrenia,^12^ epilepsy,^13^ and, as pivotally relevant in our context, focal brain lesions in general and stroke in particular. In fact, the importance of hub regions in stroke has been investigated in various ways: Variations related to the studied outcome or stroke-induced impairment, i.e., for example global functional outcome^16,17^ or more specific language impairment.^20^ Furthermore, timepoints were varied, i.e., from acute to chronic ones,^19,33^ and importantly, definitions of “hubness” differed, i.e., different measures decided about whether a brain region was considered central or not.^16,17,18,19,20,21^ Altogether, all of these innovations in qualitative and quantitative approaches in previous stroke studies generated valuable insights. They collectively point to the importance of hub regions for stroke outcome. However, most of these studies investigated the relevance of connectomic information in very circumscribed frameworks, heavily built upon *a priori* chosen hub regions and only tested small sets of hypotheses in small to medium-sized datasets, on occasion resulting in conflicting results. Warren and colleagues, for example, explicitly tested whether hubs identified based on *two* specific resting-state fMRI and hence grey matter-focused measures explained severe and widespread cognitive deficits after cerebral lesions (i.e., high system density/participation coefficients versus high degree centrality).^33^ In contrast, Reber and colleagues compared the associations of *two* grey and white matter-based measures with cognitive impairment after focal brain lesions (i.e., high participation coefficient versus high edge density).^19^ Taken together, our novel methodological framework complements these previous approaches by alleviating the necessity of defining a narrow, specific alternative hypothesis. Rather, our framework allows testing of the rich club solution against a great variety of random brain region constellations, independent of any *a priori* formulated connectomic measures. This more agnostic approach enhances the validity and reliability of rich club involvement further, especially given our comparably large sample size. This sample size aspect allows good whole-brain coverage (**Figure 2**) and grants the possibility that the rich club combination, as well as other kinds of constellations can be tested in a meaningful way (c.f., **Supplementary Table 1** for an explicit count of how often each rich club brain region was affected).

### Clinical implications of lesions in rich club regions and the question *why*?

Our analyses indeed confirmed a predominant and spatially unique role of rich club regions in explaining modified Rankin Scale (mRS)-based unfavorable outcome. In the following, we will break down the specifics, implications and potential explanatory factors of this finding. The modified Rankin Scale represents a very global assessment of stroke sequelae (0: no symptoms, 2: slight disbility, 3: moderate disability, 6: death).^34,35^ With an mRS>2 as cut-off, our distinction between favorable and unfavorable outcome reflected the change from slight to moderate disability and the ability to look after daily activities independently versus requiring some help. Hence, while being coarse-grained, this favorable versus unfavorable distinction captured appreciable clinical, subjectively meaningful effects on patients’ lives. Essentially, the combination of this ascertained real-world value and the ease of its collection has motivated the widespread reliance on the mRS as primary, FDA-^36^ and NINDS-endorsed^37^ endpoint in the majority of acute stroke treatment trials.^38,39,40^

Altogether, the relevance of our chosen outcome underscores the salience of our findings: Given the detrimental effect of stroke lesions specifically affecting rich club regions, there is, at the same time, the promise that rescuing rich club region tissue could enhance outcomes in clinically significant ways. More concretely, our findings suggest that, when in doubt and weighing treatment options, offering acute thrombolytic treatment or endovascular thrombectomy could be particularly impactful in case of rich club lesions.

Our study also motivates the investigation of several new follow-up questions to render treatment recommendations to be even more specific. Future studies will be necessary to increase the level of detail further, both with respect to behavioral, as well as brain measures. For example, it will be crucial to carve out the importance of *individual* rich club regions: Do all regions affect the outcome equally or to varying degrees? Do several of them have to be affected for noticeable sequelae or is one region sufficient? Is there a hemispheric bias? In addition, is it only the direct injury to rich club regions that is detrimental to outcomes, or also indirect ties to the lesions? Such evaluations of indirect effects will be possible thanks to more recently developed techniques to estimate structural^41,42^ and functional lesion connectivity.^43,44^ In sum, these additional pieces of information will be of particular relevance, as the rate of affection and therapeutic accessibility differ for the individual rich club regions. In view of the anatomy of the human vasculature and classic locations of vessel occlusions, there are naturally more patients experiencing a stroke that affects the putamen or thalamus than superior frontal or precuneous cortices.^45^ For example, in our sample ∼200 and ∼300 patients had lesions affecting the thalamus or putamen, respectively, and only ∼30 and ∼50 for the superior frontal or precuneous cortex (c.f., **Supplementary Table 1** for an overview). Hence, the clinical actionability critically hinges upon the importance of rich club regions, especially given the amenability of more proximal vessel occlusions to endovascular thrombectomy. Optimally, all of these analyses will be conducted in combination with methodological approaches ensuring spatial specificity, as showcased here. In addition, it will be promising to continue exploring the specifics of rich club lesion links to acute and chronic impairments in individual domains, such as sensory, motor, visual, cognitive, their combination (e.g., cognitive and motor impairments^46^) and domain-specific *recovery* trajectories.^38^ In an ideal scenario, various behavioral domains would be evaluated in the same stroke sample to allow for direct comparisons. Are lesions in rich club regions linked to differing domains with equal or varying strength? Are they particularly crucial for the actual recovery, independent of the initial impairment?

Previous work suggests that rich club regions might be particularly susceptible to brain disease given their unique properties, such as exceptionally high baseline activities and associated metabolic needs,^7,47,48^ longer-distance connections^49^ and a high proportion of shortest paths passing through^7^ (c.f., ^14^ for an excellent overview). In fact, empirical evaluations emphasize the rich club region involvement in neuropsychiatric disease, with schizophrenia and Alzheimer’s disease exhibiting the most pronounced associations.^50^ In case of Alzheimer’s disease, it has been hypothesized that it is precisely their higher baseline activity that may underlie the observable preferential accumulation of amyloid-beta in hub regions.^11^ Rich club region lesion status was shown to be informative about the *acute* symptom burden post-stroke.^16,17^ The apparent link of hub region affection and global cognitive decline in Alzheimer’s disease however raises the question whether stroke ischemia-induced disturbances of hub region integrity could reduce the capability to *recover* in general. Independent of the acute degree of impairment and specifically affected domain, patients may have a greater potential to recover any kind of function in the case of unaffected rich club regions.

### Sex-specific aspects of rich club relevance

Furthermore, our data are indicative of a female-pronounced rich club effect on functional outcomes. If lesioned in a female brain, rich club regions increased the likelihood of unfavorable outcomes substantially more than if lesioned in a male brain.

Sex differences in the human brain represent a delicate and highly debated topic. A recent comprehensive review on neuroimaging-based cerebral sex differences concluded that the human brain was “not ‘sexually dimorphic’” : According to the author’s evaluation, sex/gender explained only 1% in total variance of structural differences once brain size was taken into account (brain size, in turn, is consistently found to be ∼12% higher in males^51^).^52^ In response, others^53^ have argued that sex differences with small effect sizes, while not representing “sexual dimorphisms”, may still entail meaningful behavioral consequences, for example, if affecting repeated events.^54^ Furthermore, it may be worth considering that, even if men and women categorically only differed in their brain sizes, it might be a difference of high clinical relevance: Previous research suggests that outcomes are more favorable in case of larger brain volumes.^55^

More fine-grained analyses of structural connectivity in healthy participants have demonstrated enhanced within-hemisphere connectivity and modularity in men, in contrast to higher between-hemisphere connectivity and cross-module participation in women.^56^ With respect to functional connectivity, a large-scale study in ∼5.000 UK Biobank participants detailed stronger functional connectivity in unimodal sensorimotor areas in men, while women were characterized by stronger connectivity in the default mode network (DMN).^57^ Given the large overlap of brain regions thought to be part of the rich club on the one hand and the DMN on the other hand, this female-specific enhanced DMN connectivity could contribute to explain the greater vulnerability to rich club lesions in women. Lesions in a female brain could conceivably lead to a more far-reaching impairment of whole-brain processing. Initial sex-specific lesion network mapping-based explorations of lesion pattern effects also point in the direction of more far-reaching disruptions of functional connectivity underlying the more pronounced lesion pattern effects in women.^23^

Altogether, our findings suggest pronounced sex differences in the relevance of injury to rich club regions. It is important to realize that those cerebral sex differences were apparent in relation to behavior, i.e., we investigated the interaction between rich club effects and biological sex on the functional outcome. Our stroke patients were also on average ∼30 years older than most subjects in studies of healthy participants. Such an age difference has dramatic effects on hormonal levels of both estrogen in women, as well as testosterone in men^58^ and may alter cerebral functioning via activational effects.^59^ Therefore, these two key differences could already explain why comparable rich club region effects were not observed in previous studies comparing rich club regions in male and female brains without any links to behavior or age.^60,61^ Future studies interrogating sex differences could generate novel insights by more frequently embracing some of this additional complexity. In particular, sex differences may be modified by additional factors, such as age, socioeconomic status, education, sexual orientation and sex hormones,^53^ which need to be explicitly incorporated in analytical approaches. Stroke and further neuropsychiatric diseases, with Alzheimer’s disease being a primary example, may be promising model diseases, given their intricate links to age and significantly impacted behavior.

### Strength and limitations

The current work has several strengths and limitations: First, we had access to a large stroke database, that, due to it’s multicenter character, may be representative of a common stroke trial patient population. In combination with our comprehensive Bayesian modeling and cross-validation scheme, these factors may lay the foundation for a successful subsequent generalization to new stroke samples and individual patients. However, patients had overall fairly small lesions and were relatively mildly affected by their strokes (∼73% with favorable outcomes), which warrants additional analyses in samples of more severely affected participants experiencing large lesions on average. Plausibly, findings could be even more pronounced than reported here. Information on acute treatment or the pre-stroke level of stroke was furthermore not readily available for the entire cohort. As recommended in the literature,^62^ we computed continuous values for region-wise lesion status (i.e., we calculated how many voxels were damaged per region), rather than applying a binarizing damage threshold. Nonetheless, we focused on a binary outcome – favorable versus unfavorable outcomes – and continuous, more granular and domain-specific outcome measures may facilitate even more detailed insights, as outlined above. We here adopted the definition of rich club regions based on white matter-focused and hence structural diffusion tensor imaging (DTI)-derived measures, such as the node-specific degree or strength, as put forward in the fundamental work by van Heuvel and Sporns.^6^ Alternative definitions exist, and it would have been equally valid to define “hubness” based on grey matter-focused and functional resting state fMRI data-defined measures, such as the participation coefficient.^19^ Similarly, future work could evaluate different numbers of rich club regions, as we here strictly relied on the number six, as established and employed in prior work.^6,16^

## Conclusions

Employing comprehensive Bayesian modeling techniques and permutation analyses, we here demonstrate the spatial specificity of rich club regions in their relevance for long-term stroke outcomes. Notably, this rich club effect on outcomes post-stroke was substantially more pronounced in women as compared to men. More research is needed to determine further intricacies of these rich club effects. Nevertheless, our findings suggest that taking lesions to rich club regions into account when deciding about patient-centered, individualized acute stroke treatments allows greater understanding of longer-term outcomes after stroke. Our results support the notion of intimate links between rich club region malfunction and neuropsychiatric diseases and may hold the promise to explain cerebral sex differences beyond ischemic stroke.

## Data Availability

The authors agree to make the data available to any researcher for the express purposes of reproducing the here presented results and with the explicit permission for data sharing by individual sites' institutional review boards.

## Acknowledgments

We are grateful to our colleagues at the J. Philip Kistler Stroke Research Center for valuable support and discussions. Furthermore, we are grateful to our research participants without whom this work would not have been possible.

## Supplemental materials

### Methods

#### Sample size calculation

Automatically segmented DWI-defined stroke lesions were available for 2,765 MRI-GENIE patients.^1^ A total of 1,920 (70.1%) were approved in internal quality control by two experienced raters (M.B., A.K.B.). The availability of information on covariates and ∼3 months mRS outcomes then determined the final number of included 822 patients.

#### Neuroimaging parameters

Neuroimages were obtained in 1T, 1.5T or 3T scanners (General Electric Medical Systems, Philips Medical Systems, Siemens, Toshiba, Marconi Medical Systems, Picker International, Inc.).

#### Diffusion-weighted images (DWI)

Mostly axial orientation (2727/2770 axial, 43/2770 coronal). Axial: Reconstruction matrix 256×256mm^2^ (range: 128×128mm^2^ to 432×384mm^2^), median field-of-view 230 mm (range: 200 to 420 mm), median slice thickness 5mm (range: 2 to 7mm, gaps of 0 to 3mm), median TR 4.773ms, median TE 92ms. Coronal: reconstruction matrix 256×256 mm^2^, median field-of-view 260mm, median slice thickness 5mm, median TR 8.200ms, median TE 112ms. Mostly 3 directions (range: 3 to 25). Mostly low b-value 0s/mm^2^ (range: 0 to 50s/mm^2^), high b-value 1000s/mm^2^ (range: 800 to 2000s/mm^2^).

### Full model specifications

#### Hyperpriors

*σ_ β ∼ Halfcauchy(1)*

*hyper_mu_ β ∼ Normal(µ = 0, σ = 1)*

*mu_ β*_*rich club, non-rich club*_ *∼ Normal(µ = hyper_ mu_ β, σ = 1)*_*rich club, non-rich club*_

#### Priors

*⍰ #x223C; Normal(µ = 0, σ = 1)*

*β*_*1-12; rich club*_ *∼ Normal(µ = mu_ β*_*rich club*_, *σ = σ_ β)* _*1-12; rich club*_

*β*_*1-96; non-rich club*_ *∼ Normal(µ = mu_ β*_*non-rich club*_, *σ = σ_ β)* _*1-96; non-rich club*_

_*β*__*age*_ *∼ Normal(µ = 0, σ = 1)*

*β*_*age*age*_ *∼ Normal(µ = 0, σ = 1)*

*β*_*sex*_ *∼ Normal(µ = 0, σ = 1)*

*β*_*hypertension*_ *∼ Normal(µ = 0, σ = 1)*

*β*_*diabetes*_ *∼ Normal(µ = 0, σ = 1)*

*β*_*atrial fibrillation*_ *∼ Normal(µ = 0, σ = 1)*

*β*_*coronary artery disease*_ *∼ Normal(µ = 0, σ = 1)*

*β*_*prior stroke*_ *∼ Normal(µ = 0, σ = 1)*

*β*_*smoking*_ *∼ Normal(µ = 0, σ = 1)*

*β*_*log total stroke lesion volume*_ *∼ Normal(µ = 0, σ = 1)*

#### Likelihood

*mRS_bin_est = ⍰ + β*_*j=1…12*_,_*rich club*_*\* rich club lesion volume*_*j=1…12*_,_*rich club*_ *+ + β*_*k=1…96*_,_*non-rich club*_*\* Non-rich club lesion volume*_*k=1…96*_,_*non-rich club*_ *+ β*_*age*_ *\* Age + β*_*age*age*_ *\* Age* ^*2*^ *+ β*_*sex*_ *\** Sex + *β*_*hypertension*_ * hypertension + *β*_*diabetes*_ * diabetes + *β*_*atrial fibrillation*_ * atrial fibrillation + *β*_*coronary artery disease*_ * coronary artery disease + *β*_*prior* stroke_ * prior stroke + *β*_*smoking*_ * smoking + *β*_*log total stroke lesion volume*_ * total stroke lesion volume

*Unfavorable functional outcome* ∼ *Bernoulli(p = deterministic_sigmoid(mRS_bin_est))*

**Supplementary Figure 1.**
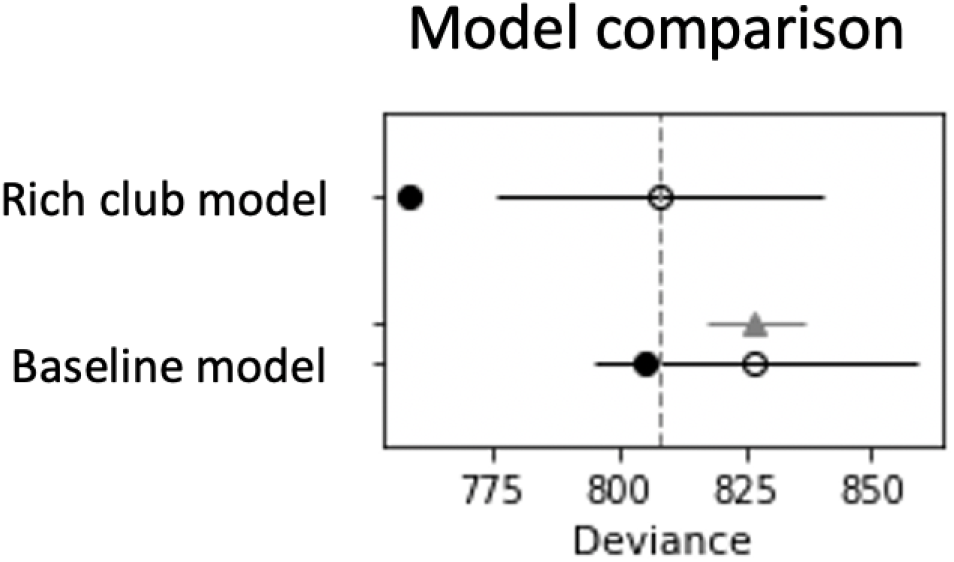
Bayesian model comparison of rich club model and covariate-based baseline model of unfavorable outcomes. The area under the curve (AUC) of the rich club model was 0.80, while the baseline model had an AUC of 0.77.

**Supplementary Table 1.** Rich club region-wise count of patients with a lesion affecting the respective rich club region. Please not that we did not perform hemisphere-specific analyses.

| Superior Frontal Cortex | Superior Parietal Cortex | Precuneous Cortex | Thalamus | Putamen | Hippocampus |
| --- | --- | --- | --- | --- | --- |
| 31 | 92 | 55 | 197 | 310 | 103 |

